# Metagenomic sequencing detects human respiratory and enteric viruses in air samples collected from congregate settings

**DOI:** 10.1101/2023.05.28.23290648

**Authors:** Nicholas R. Minor, Mitchell D. Ramuta, Miranda R. Stauss, Olivia E. Harwood, Savannah F. Brakefield, Alexandra Alberts, William C. Vuyk, Max J. Bobholz, Jenna R. Rosinski, Sydney Wolf, Madelyn Lund, Madison Mussa, Lucas J. Beversdorf, Matthew T. Aliota, Shelby L. O’Connor, David H. O’Connor

## Abstract

Innovative methods for evaluating virus risk and spread, independent of test-seeking behavior, are needed to improve routine public health surveillance, outbreak response, and pandemic preparedness. Throughout the COVID-19 pandemic, environmental surveillance strategies, including wastewater and air sampling, have been used alongside widespread individual-based SARS-CoV-2 testing programs to provide population-level data. These environmental surveillance strategies have predominantly relied on pathogen-specific detection methods to monitor viruses through space and time. However, this provides a limited picture of the virome present in an environmental sample, leaving us blind to most circulating viruses. In this study, we explore whether pathogen-agnostic deep sequencing can expand the utility of air sampling to detect many human viruses. We show that sequence-independent single-primer amplification sequencing of nucleic acids from air samples can detect common and unexpected human respiratory and enteric viruses, including influenza virus type A and C, respiratory syncytial virus, human coronaviruses, rhinovirus, SARS-CoV-2, rotavirus, mamastrovirus, and astrovirus.

## Introduction

As of September 23rd, 2023, over 70 million SARS-CoV-2 diagnostic tests have been performed in the United States ^1^. Deploying individual testing programs at this scale is extraordinarily expensive and resource-intensive, and has become increasingly unsustainable in most jurisdictions. New strategies are needed for monitoring the spread and evolution of pathogens without relying on this widespread individual testing.

Environmental surveillance, be it through wastewater or air, shows promise for meeting this need. Without relying on individual testing, environmental surveillance has already enabled public health officials to rapidly assess infection risk in congregate settings and in communities writ large^2–5^. In the COVID-19 pandemic, the majority of environmental sampling for surveillance purposes has been through wastewater. However, air sampling has several advantages that make it complementary or even preferable to wastewater in certain settings. For example, air samplers are portable and intrinsically hyperlocal; they can be moved between individual rooms, installed into HVAC systems, and deployed densely in more open areas like airport terminals. Additionally, wastewater sampling may be unfeasible in some areas, such as rural areas where wastewater is disposed of in septic systems. Perhaps most importantly, many human viruses transmit predominantly through aerosols or respiratory droplets, which is exactly what air samplers are designed to collect. Air sampling makes it possible to identify a wide variety of aerosolized viruses while they are in the process of potentially spreading between hosts ^6–9^.

However, few studies or real-world air sampler deployments have taken advantage of this bioaerosol diversity. Doing so would require the development of virus-agnostic, metagenomic detection methods, which, when combined with air sampling, could expand surveillance to any airborne virus. Early contributions in this area, e.g. ^10–14^, demonstrate that it is possible to detect human viruses in various settings. However, this detection comes with a variety of technical challenges ^15^. First, the relative abundance of aerosolized human viruses in the above studies’ air samplers was extremely low. For example, Prussin et al. 2019 characterized airborne viral communities in a daycare center’s HVAC system over one year ^11^. Over that year, commonly circulating human viruses accounted for less than 0.005% of the total genetic material, with the majority of total virus sequences coming from bacteriophages and plant-associated viruses. A second challenge is defining which portion of each viral genome to use for classification. Bacteria and fungi have universal genetic marker regions (16S and internal transcribed spacer (ITS) ribosomal RNA, respectively) that are used for sequencing and classification. In contrast, there is no single genetic marker shared across the many viruses that could be present in the air. This leaves unbiased amplification of human virus genetic material as the best option for detecting many, potentially underappreciated viruses in the air ^16^. One especially promising method is sequence-independent single-primer amplification (SISPA) ^17^, which has been used to detect a wide range of viruses in clinical samples ^18^.

In 2021, we reported on the characterization of SARS-CoV-2 and other respiratory viruses in air samples collected from congregate settings^19^. We also used a semi-quantitative PCR assay in that study to detect 40 other pathogens, demonstrating that air samples could be used to monitor the variety of pathogens that may be present in the spaces around us. Here, we use SISPA to detect an even broader array of human RNA viruses from air collected in congregate settings. Understanding human viruses in built environments’ air may help elucidate illness trends in communities over time. This approach could enhance air sampling as a tool for public health virus surveillance and preparedness against many emerging and re-emerging viruses.

## Results

### Study design

From July 2021 to December 2022, we deployed active air samplers in several community settings in the Upper Midwestern states of Wisconsin and Minnesota for routine pathogen monitoring. Thermo Fisher AerosolSense Samplers were used to collect air samples from high-traffic areas in several different congregate settings, including a preschool, campus athletic facility, emergency housing facility, brewery taproom, household, and five K-12 schools. Air samples were collected at weekly and twice-weekly intervals as previously described ^19^. To demonstrate the feasibility of pathogen-agnostic sequencing to detect human viruses captured in air samples in real-world settings, we analyzed a total of 22 air samples across the 10 congregate settings (Table 1). We also processed three air sample filter substrates from unused AerosolSense cartridges, as no-template controls. Viral RNA was extracted from air samples, and complementary DNA (cDNA) was prepared using sequence-independent single primer amplification (SISPA) for Oxford Nanopore deep sequencing and metagenomic analysis. Sequencing reads were filtered for host and reagent contaminants and mapped to 835 human-associated viral reference sequences from NCBI to look for common circulating RNA and DNA viruses (available on GitHub at https://github.com/dholab/air-metagenomics/blob/main/resources/ncbi_hu-man_virus_refseq_20221011.masked.fasta).

**Table 1.** Human RNA viruses detected in air samples through deep sequencing with the Oxford Nanopore PromethION.

| Air sample ID | Location | Start | End | Total number of reads | Pathogen | Number of mapped reads | Percent genome coverage |
| --- | --- | --- | --- | --- | --- | --- | --- |
| HC211129 | Brewery Taproom | 11.22.21 | 11.29.21 | 4,113,547 | Rotavirus A | 276 | 57.2% |
| AE000010795B42 | Emergency housing shelter | 12.21.21 | 1.7.22 | 4,898,090 | SARS-CoV-2 | 266 | 6.5% |
| AE00001004492C | Campus athletic facility | 12.22.21 | 12.23.21 | 4,566,986 | Respiratory syncytial virus B | 182 | 16.7% |
| AE0000100A9F46 | Preschool | 1.5.22 | 1.19.22 | 7,037,819 | Human astrovirus | 70 | 10.6% |
| AE0000100A8839 | Preschool | 1.26.22 | 2.1.22 | 4,770,197 | Human astrovirus | 207 | 66.3% |
|  |  |  |  |  | Respiratory syncytial virus A | 220 | 68.9% |
|  |  |  |  |  | Mamastrovirus | 41 | 26.2% |
|  |  |  |  |  | Influenza C virus | 15 | 9.2% |
| AE0000100A8B3C | Preschool | 2.1.22 | 2.8.22 | 4,022,365 | Influenza C virus | 1,826 | 95.0% |
|  |  |  |  |  | SARS-CoV-2 | 220 | 46.7% |
|  |  |  |  |  | Human rhinovirus | 102 | 68.6% |
|  |  |  |  |  | Human astrovirus | 48 | 40.2% |
|  |  |  |  |  | Mamastrovirus | 47 | 36.8% |
| AE0000100A9532 | Preschool | 2.23.22 | 3.1.22 | 8,252,402 | Human coronavirus NL63 | 921 | 19.3% |
|  |  |  |  |  | Influenza C virus | 884 | 38.83% |
| AE0000100C3430 | Elementary school 1 | 4.11.22 | 4.19.22 | 3,205,370 | Human coronavirus 229E | 8 | 13.2% |
| AE0000100CA130 | Elementary school 2 | 3.7.22 | 3.14.22 | 4,606,042 | Human astrovirus | 12 | 46.2% |
|  |  |  |  |  | Human Coronavirus NL63 | 7 | 11% |
|  |  |  |  |  | Human coronavirus 229E | 4 | 6% |
| AE0000100CD638 | Elementary school 2 | 4.4.22 | 4.18.22 | 3,945,143 | Rotavirus A | 50 | 14.93% |
| AE0000100CA232 | Elementary school 2 | 4.18.22 | 5.2.22 | 3,212,852 | Human coronavirus 229E | 11 | 5.8% |
| AE0000100C7A40 | Elementary school 2 | 5.2.22 | 5.9.22 | 4,958,920 | NA | NA | NA |
| AE0000100C702C | Elementary school 2 | 5.9.22 | 5.16.22 | 3,788,598 | Rotavirus A | 31 | 37.8% |
| AE0000100CC536 | Elementary school 2 | 5.16.22 | 5.23.22 | 3,920,258 | Rotavirus A | 105 | 40.7% |
| AE0000100CC63D | Elementary school 2 | 5.23.22 | 5.31.22 | 4,836,345 | Rotavirus A | 40 | 7.2% |
|  |  |  |  |  | Simian Agent 10 | 7 | 8.9% |
| AE000010014930 | Elementary school 2 | 11.10.22 | 11.14.22 | 8,999,110 | Human coronavirus HKU1 | 5 | 2.2% |
| AE000010011724 | Elementary school 2 | 11.22.22 | 12.1.22 | 4,946,530 | Rotavirus A | 3 | 10.2% |
| AE00001073172E | Elementary school 3 | 12.5.22 | 12.8.22 | 7560971 | Human coronavirus HKU1 | 113 | 36.1% |
| AE00000FEC714A | Middle school | 10.31.22 | 11.3.22 | 7,421,046 | NA | NA | NA |
| AE00000FEC2245 | Middle school | 12.5.22 | 12.8.22 | 9,380,984 | NA | NA | NA |
| AE0000106DC33E | High school | 11.28.22 | 12.1.22 | 6,669,497 | Influenza A virus | 30 | 10.7% |
| AE0000100DE93E | Household | 4.22.22 | 4.26.22 | 6,810,106 | Human rhinovirus | 17 | 17.8% |
| NTC_27269_1 | No template control | NA | NA | 26,833 | NA | NA | NA |
| NTC2_27269_2 | No template control | NA | NA | 5,158 | NA | NA | NA |
| NTC_28210 | No template control | NA | NA | 253,439 | NA | NA | NA |

Percent genome coverage was calculated by summing the total number of basepairs with any amount of read coverage divided by the total number of basepairs for the given virus target in the RefSeq file. Dates are listed as MM.DD.YY. Abbreviations: NA, not applicable.

### Detection of human respiratory and enteric viruses

Deep sequencing allowed us to detect human viruses in 19 out of 22 (86%) air samples. No human viruses were detected in any of the no-template controls. We define a detection of a virus “hit” as two or more reads mapping to the viral reference sequence in two or more non-overlapping genomic regions. By this definition, we detected a total of 13 human RNA viruses in air samples (Table 1). Several of these viruses are associated with frequent and seasonal respiratory illnesses that cause a burden on the healthcare system, including influenza virus type A and C, respiratory syncytial virus subtypes A and B, human coronaviruses (NL63, HKU1, and 229E), rhinovirus, and SARS-CoV-2 (Figure 1). We also detected human viruses associated with enteric disease, including rotavirus, human astrovirus, and mamastrovirus, in ten of the 22 (45%) air samples in this study.

**Figure 1.**
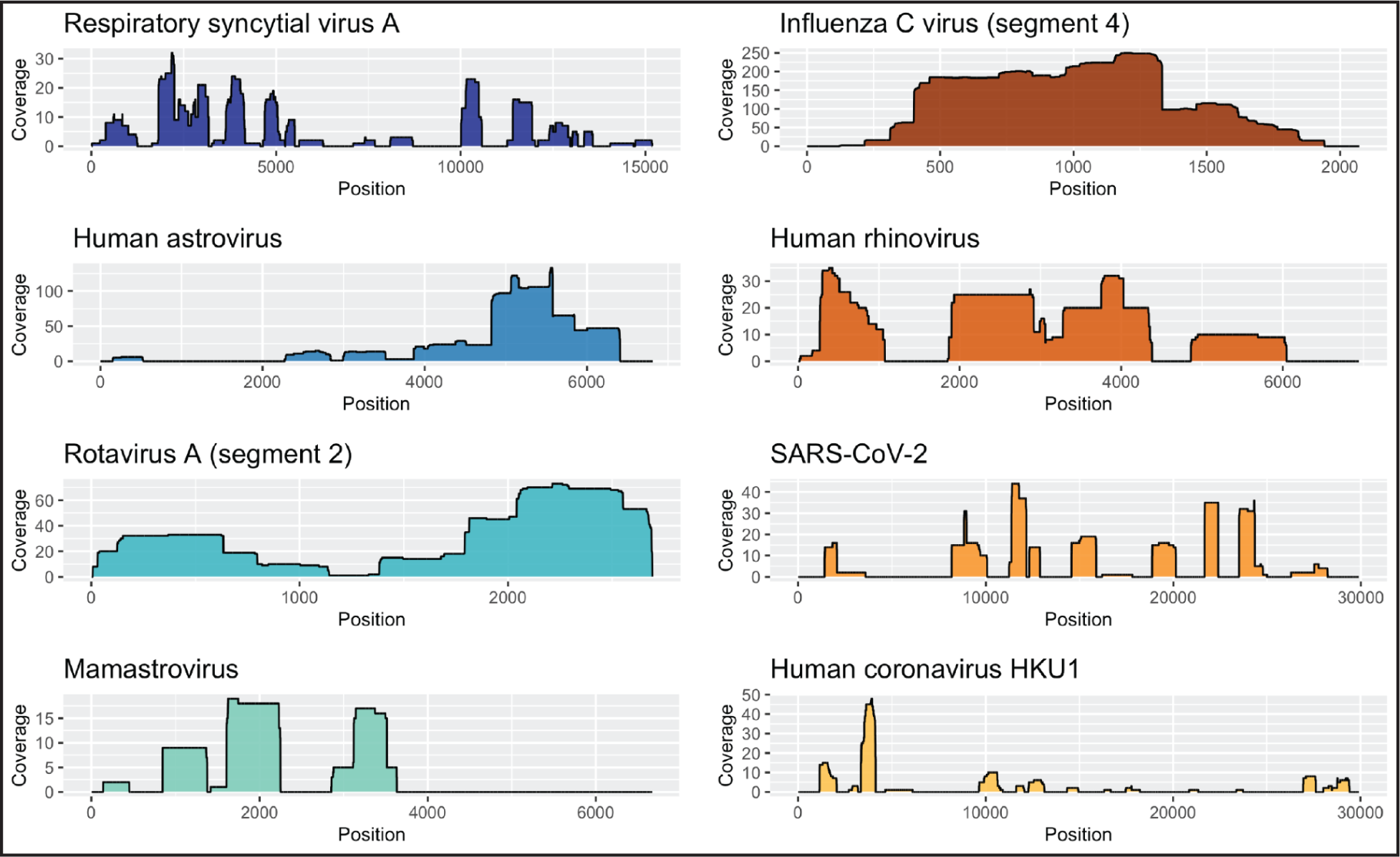
Human respiratory and enteric viruses detected in air samples by SISPA amplification and Oxford Nanopore sequencing. Genome coverage plots showing read depth across eight human RNA viruses detected in air samples. The depth of coverage is shown on the y-axis, and the genome position is shown on the x-axis. The scale of the y-axis varies between plots. Coverage plots were created using ggplot2 (3.4.1) using a custom vR script (4.2.3) in RStudio (2023.03.0+386).

### Characterizing influenza C virus lineage in a preschool air sample

A preschool air sample from the week of February 1st showed comprehensive coverage of influenza C virus (ICV), with reads mapping to all seven gene segments (supplementary data 1). This included hemagglutinin-esterase (HE), each of the genes encoding proteins for the polymerase complex (PB2, PB1, and P3), nucleoprotein (NP), matrix (M), and nonstructural protein (NS) (supplementary data 1). ICV is an understudied respiratory virus, with a total of 2,475 ICV sequences available in NCBI Genbank (taxid:11552) and only 134 ICV sequences submitted from the United States in the 21st century. To contribute more data on this understudied virus, we used the remaining SISPA-prepared cDNA from the February 1st preschool sample to perform an additional sequencing run with the Oxford Nanopore GridION. This increased the sample’s depth of coverage across the ICV genome compared to the initial Oxford Nanopore PromethION run, where the flow cell was shared with many samples. We then used the GridION reads to create consensus sequences for each gene segment, which enabled us to perform a Tamura-Nei neighbor-joining phylogenetic analysis that compared the February 1st preschool ICV with 45 other ICV viruses from GenBank (supplementary data 1). Our phylogeny supported six genetic lineages for the HE gene and two for all other gene segments (Figure 2; supplementary figure 1). HE grouped with the C/Kanagawa/1/76 lineage. PB2, PB1, M, and NS grouped with the C/Yamagata/81 lineage. P3 and NP group with C/Mississippi/80 lineage. This particular reassortment clusters closely with the influenza C virus C/Scotland/7382/2007, which was previously identified by Smith et al. (Figure 2; supplementary figure 1) ^20^.

**Figure 2.**
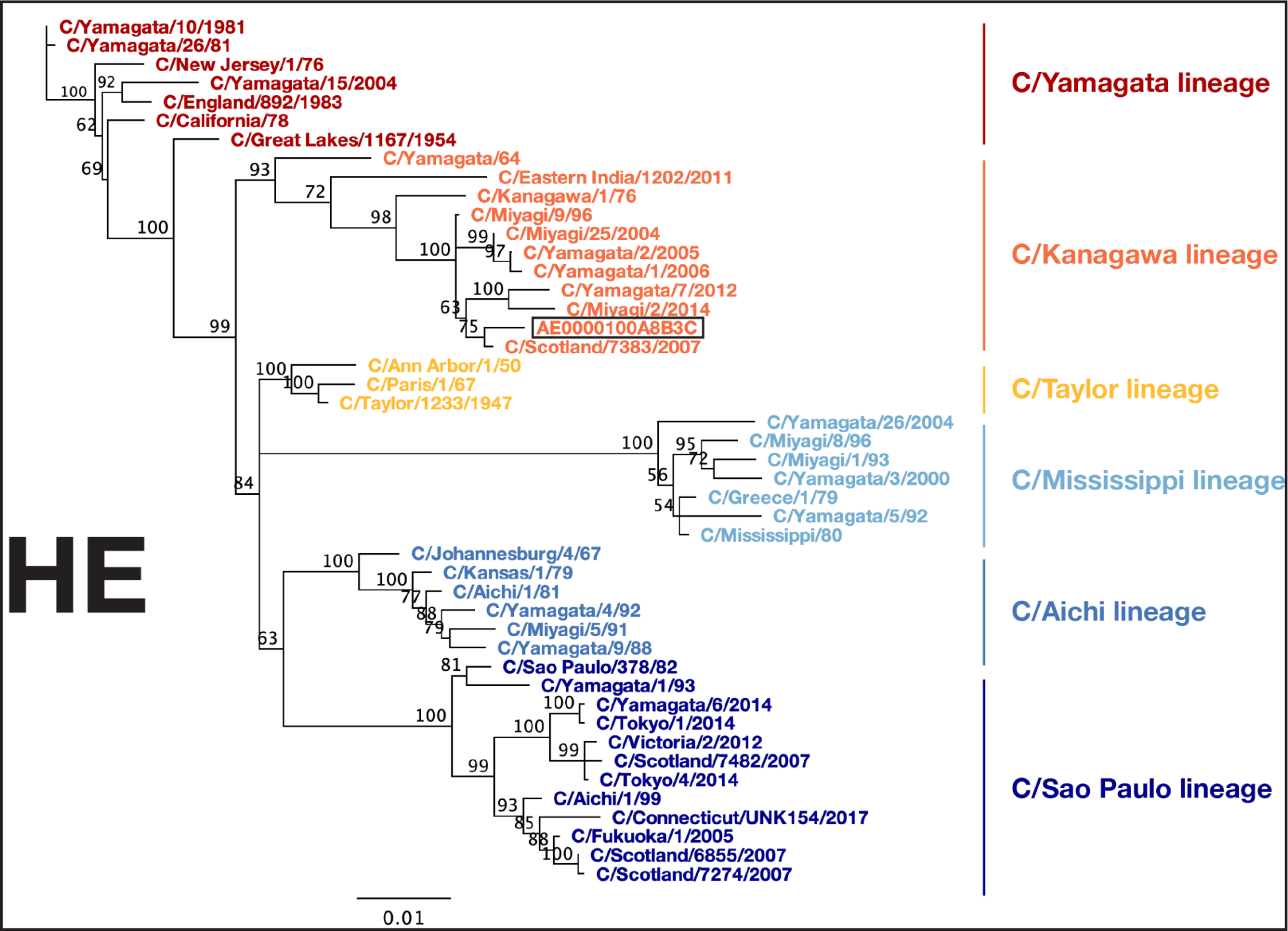
Phylogenetic analysis of influenza C virus hemagglutinin-esterase gene. Phylogenetic trees for influenza C virus hemagglutinin-esterase (HE) gene segment. Nucleotide sequences were aligned using MUSCLE (5.1). The phylogenetic tree was constructed with the Geneious Tree Builder (2023.0.4) using the Neighbor-joining method and Tamura-Nei model with 100 bootstrapped replicates. Numbers above the branches indicate the bootstrap values with 100 replicates. ICV strain names are listed at the end of branches. ICV strains belonging to the C/Sao Paulo lineage are represented in dark blue, C/Aichi lineage in blue, C/Mississippi lineage in light blue, C/Taylor lineage in yellow, C/Kanagwa lineage in orange, C/Yamagata lineage in red.

### Longitudinal detection of human viruses in a preschool

Metagenomic analysis of air samples longitudinally collected from congregate settings can provide insight into changes in the prevalence of pathogens over time. These data could provide public health authorities valuable information to improve routine pathogen surveillance programs and outbreak investigations. To track the prevalence of viral genetic material from ICV and other human viruses in a preschool, we analyzed four air samples that were longitudinally collected from January 5, 2022, to March 1, 2022. ICV was first detected in an air sample collected from January 26th to February 1st, 2022. Viral reads in this sample mapped to three out of the seven gene segments including HE, PB2, and NS. Two air samples collected after February 1st, 2022, also contained reads that mapped to several ICV gene segments. ICV genetic material was detected at the highest abundance in air sample AE0000100A8B3C collected from February 1st to the 8th, with reads mapping to all seven gene segments as described above (Table 1; supplementary data 1). Viral reads mapping to five of the seven gene segments, including PB2, PB1, P3, HE, and NP, were detected in an air sample collected from February 23rd to March 1st.

### Detection of SARS-CoV-2 in RT-PCR-positive air samples

To explore whether metagenomic sequencing can detect a human virus that is known to be present in an air sample, we sequenced air samples with known SARS-CoV-2 status. Each AerosolSense cartridge comes with two filter substrates. One filter substrate from each air sample was tested by reverse transcription PCR (RT-PCR) to determine its SARS-CoV-2 status. The other substrate was eluted in 500ul of PBS and stored at -80℃ until it was processed for sequencing. Several different RT-PCR assays were used on samples included in this study, depending on when and where they were collected, as previously described ^19^. Cut-off values used for determining if an air sample was positive, inconclusive, or negative for SARS-CoV-2 are described in the methods section.

SISPA sequencing was able to detect SARS-CoV-2 reads in two out of 15 (13.3%) air cartridges that were positive for SARS-CoV-2 by the more sensitive RT-PCR assays (Table 1; supplementary data 1). The percent of genome coverage varied between the two samples (6.5% and 46.7%). No SARS-CoV-2 reads were observed in any of the samples that were negative or inconclusive for SARS-CoV-2 by RT-PCR testing or with no template controls (supplementary data 1). An inconclusive result was defined as a sample with only amplification in one of the PCR targets. These data suggest that, unsurprisingly, SISPA sequencing is not as sensitive as RT-PCR for detecting viral genomic material captured in the air samples.

## Discussion

In this study, we used air samples and metagenomic sequencing to detect human RNA viruses in a variety of congregate settings. Specifically, our results show that active air sampling, SISPA library preparation, long-read Oxford Nanopore sequencing, and metagenomic bioinformatics can be used to detect both common and lesser-known human viruses. Because air samplers collect bioaerosols produced when infected individuals breathe, sneeze, cough, or talk, the majority of detected viruses were respiratory. However, we also detected enteric viruses that are transmitted through the fecal-oral route^21,22^. Several studies have previously used air sampling to detect enteric viruses in congregate settings, including a daycare, a wastewater treatment facility, and a hospital^11,14,23^. The detection of respiratory viruses in wastewater, and enteric viruses in air samples, creates future opportunities for integration of clinical, wastewater, and air sampler data from the same geographical location to obtain a more comprehensive understanding of viral spread within communities.

The most unexpected virus we detected was Influenza C Virus (ICV) in a preschool. SISPA and Oxford Nanopore sequencing allowed us to classify the viral lineages of all seven gene segments of ICV. ICV is a lesser-studied influenza virus that is often excluded from routine respiratory pathogen surveillance programs, which highlights one important limitation of virus-specific surveillance. Despite previous studies having shown a high seroprevalence of ICV in children increasing in age, it suggests that this is a common yet under-ascertained cause of respiratory illness ^24–26^, with an epidemiology that remains poorly known^26,27^. Our ICV results highlight the potential of using air sample networks to detect viruses that previously had limited awareness. ICV was historically very difficult to detect with cell culture techniques because it causes weak cytopathic effects ^24^, which may lead to an underestimation of ICV prevalence. It will be interesting to see how much more often viruses like ICV are detected as virus-agnostic environmental surveillance becomes more prevalent.

A potential strength of regular air sample collection, processing, and analysis is characterizing outbreaks longitudinally. For example, we detected the same ICV in the same preschool at four instances, with viral read count rising and falling through time. One possible explanation for this pattern is that it reflects airborne viral RNA load rising and falling over the course of the source infection(s), though we do not have the data to assess host viral load itself. We detect a similar pattern in longitudinal pre-school air sampling for astrovirus, which showed an increase and trailing off of sequencing reads between samples. These results suggest that air sampling can be used to characterize outbreaks in real time. In extended outbreaks, it could also be used to provide early sequencing results, which could then be used to design primers for more sensitive amplification of viruses in the air.

All said, we caution that our study does not include a baseline clinical knowledge of all viruses that were present in the settings we sampled. More study will be needed to understand the relationship between air-sampler-derived read counts and airborne viral loads, to determine when a viral load is too low to be detected by our methods, and to rigorously assess the sensitivity and specificity of this approach to pathogen monitoring. However, our SARS-CoV-2 detection results suggest that improving sensitivity may be the best place to start optimizations: while we never detected SARS-CoV-2 in the absence of known infections (no false positives), we failed to detect it in 85% of the cases where an air sample tested positive by a more sensitive qPCR assay.

Improvements in bioaerosol collection, nucleic acid library preparation, sequencing technology, and pathogen-agnostic bioinformatics could all significantly improve the detection of human pathogens with air samplers. While SISPA and Oxford Nanopore sequencing enabled us to detect portions of a variety of viral genomes, we inevitably missed additional viruses that were present in the air. Air samples contain high amounts of human, animal, and microbial ribosomal RNA (rRNA), likely associated with airborne microbes and host cells transported on dust particles ^28^. Several studies have shown that rRNA depletion can improve the sensitivity of unbiased sequencing techniques for recovering human RNA viruses from different modalities ^29^, and should be considered for use with air samples. Alternatively, probe-based target capture methods, where nucleic acids eluted from the air cartridge are only retained if they are a reverse complement of a probe sequence^30^, could be used to enrich viral target sequences. Of note, enriching for a predetermined panel of viruses would make this a multi-pathogen method, not a pathogen-agnostic method^29^. Even still, the line between multi-pathogen and pathogen-agnostic is increasingly blurred; commercial kits are available that contain probes for more than 3,000 different viruses, including those with ssRNA, dsRNA, dsDNA, and ssDNA genomes. Truly pathogen-agnostic methods, such as SMART-9N, are also becoming available^31^. These kits have been used with several different sample types to detect common and uncommon viruses in human and animal specimens (nasal swabs and plasma), mosquitoes, and wastewater ^32–35^ Ribosomal RNA depletion and target enrichment both show great promise for enabling air sample networks to screen for pathogens with low abundance in the air.

Rapidly evolving sequencing technology could also play a role in improving the efficacy of air sampler networks. Sequencing workflows need to be optimized to be high-throughput, cost effective, and have rapid result turnaround for widespread use with air surveillance programs. In this study we ran two Oxford Nanopore sequencing runs on the PromethION 24. The runs multiplexed 16 and 9 samples and had an output of 85 and 67 Gigabases per flow cell, respectively. The manufacturer estimates a maximal output of 290 Gigabases per flow cell when using newer sequencing kit chemistries ^36^.

Improving the sequencing yield to 200 Gigabases could allow for multiplexing up to 32 air samples, while maintaining an average of 6 million Gigabases per air sample. This could make sequencing more cost-effective while maintaining a similar per sample output obtained in this study. Additionally, Oxford Nanopore sequencing enables real-time processing of sequencing data. This could help decrease the turnaround time, as a stream of data will become available as soon as sequencing begins instead of after 72 hours of sequencing has completed. This rapid result turnaround time could be beneficial during outbreak response, when real-time data is essential.

The COVID-19 pandemic sparked widespread interest in environmental surveillance strategies for improving pandemic preparedness and outbreak response. While wastewater surveillance has many benefits, air surveillance via active air samplers is more mobile, which makes it easy to quickly deploy air sampling networks in settings of interest such as health clinics, airplanes, ports of entry, public transit, farms, K-12 schools, long-term care facilities, emergency housing facilities, or any other setting where people from many places congregate ^37–39^. Highlighting this potential application, Mellon et al. recently deployed AerosolSense samplers in an outpatient clinic for patients suspected of mpox infection to look for mpox virus in the air during the 2022 mpox public health emergency of international concern ^40^. Given the portability and flexibility of deployment, air sampling and detection of viral nucleic acids from these samples could become a cornerstone of agile public health responses to viral outbreaks in the near future.

Recent advances in metagenomic sequencing technologies have increased efforts to study microbial communities in built environments. This study demonstrates that metagenomic sequencing approaches paired with air sampling can be used to detect human respiratory and enteric viruses of public health importance in real-world settings. With continual technological improvements and laboratory optimizations, the general framework put forth here could provide a rapid means of monitoring viruses without relying on test-seeking behavior or pathogen-specific assays.

## Methods

### Ethics Statement

Our study does not evaluate the effectiveness of air samplers for diagnosing individuals for COVID-19 or other illnesses, nor does it collect samples directly from individuals. Therefore, it does not constitute human subjects research.

### Air sample collection and processing

AerosolSense instruments (Thermo Fisher Scientific) were installed in a variety of indoor congregate settings to collect bioaerosols for pathogen surveillance from December 2021 to December 2023. AerosolSense instruments were placed on flat surfaces 1-1.5 meters off the ground in high-traffic areas of an athletics training facility, preschool, emergency housing facility, brewery taproom, and five K-12 schools in the Upper Midwestern States of Wisconsin and Minnesota. Air samples were collected using AerosolSense cartridges (Thermo Fisher Scientific) according to the manufacturer’s instructions. The iOS and Android Askidd mobile app was used to collect air cartridge metadata and upload it to a centralized Labkey database, as previously described in Ramuta et al ^19^. After the air samples were removed from the instruments, they were transferred to the lab for further processing. Two air sample substrates were removed from each of the AerosolSense cartridges using sterile forceps to place them in two separate 1.5 mL tubes containing 500 μL of PBS. The tubes were vortexed for 20 seconds, centrifuged for 30 seconds, and stored at -80℃ until RNA extraction and complementary DNA (cDNA) preparation.

### Air sample total nucleic acid extraction and concentration

Total nucleic acids were extracted from air samples using the Maxwell 48 Viral Total Nucleic Acid Purification Kit (Promega) according to the manufacturer’s recommendations. Briefly, 300 μL of air sample eluate was added to a 1.5 μL tube containing 300 μL of lysis buffer and 30 μL of Proteinase K. An unused air cartridge was processed with each Maxwell run to be used as a no-template control. The reaction mix was vortexed for 10 seconds and incubated at 56℃ for 10 minutes. Following the incubation, the tubes were centrifuged for 1 minute. Then, 630 μL of the reaction mix was added to the Maxwell 48 cartridges, which were loaded into a Maxwell 48 instrument and processed with the Viral Total Nucleic Acid program. Nucleic acids were eluted in a final volume of 50 μL of nuclease-free water. To clean and concentrate the viral RNA, 30 μL of extracted total nucleic acids were treated with TURBO DNase (Thermo Fisher Scientific) and concentrated to 10 μL with the RNA Clean & Concentrator-5 kit (Zymo Research) according to the manufacturer’s protocols.

### Air sample sequencing

A modified sequence-independent single primer amplification (SISPA) approach previously described by Kafetzopoulou et al. was used to generate cDNA from the air samples ^18,41^. First, 1 μL of Primer A (Table 2) was added to 4 μL of concentrated viral RNA and incubated in a thermocycler at 65℃ for 5 minutes, followed by 4℃ for 5 minutes. To perform reverse transcription, 5 µL of Superscript™ IV (SSIV) First-Strand Synthesis System (Invitrogen) master mix (1 µL of dNTP (10mM), 1 µL of nuclease-free water, 0.5 µL of DTT (0.1 M), 2 µL of 5X RT buffer, and 0.5 µL of SSIV RT) was added to the reaction mix and incubated in a thermocycler at 42℃ for 10 minutes. To perform second-strand cDNA synthesis, 5 µL of Sequenase Version 2.0 DNA polymerase (Thermo Fisher Scientific) master mix (3.85 µL of nuclease-free water, 1 µL of 5X Sequenase reaction buffer, and 0.15 µL of Sequence enzyme) was added to the reaction mix and incubated at 37℃ for 8 minutes. After the incubation, 0.45 µL of the Sequenase dilution buffer and 0.15 µL of Sequenase were added to the reaction mix and incubated at 37℃ for 8 minutes. To amplify the randomly primed cDNA, 5 µL of the cDNA was added to 45 µL of the Primer B reaction mix (5 µL of AccuTaq LA 10x buffer, 2.5 µL of dNTP (10mM), 1 µL of DMSO, 0.5 µL of AccuTaq LA DNA polymerase, 35 µL of nuclease-free water, and 1 µL of Primer B (Table 2)). The following thermocycler conditions were used to amplify the cDNA: 98℃ for 30 seconds, 30 cycles (94°C for 15 seconds, 50℃ for 20 seconds, and 68℃ for 2 minutes), and 68℃ for 10 minutes. The amplified PCR product was purified using a 1:1 ratio of AMPure XP beads (Beckman Coulter) and eluted in 25 µL of nuclease-free water. The purified PCR products were quantified with the Qubit dsDNA high-sensitivity kit (Invitrogen).

**Table 2.**
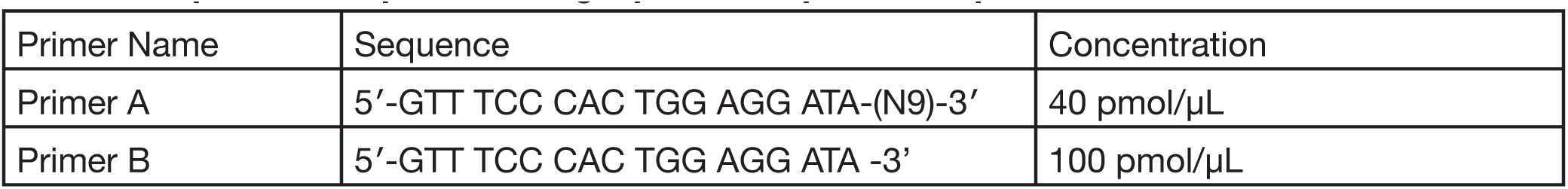
Sequence independent single primer amplification primers.

### Oxford Nanopore sequencing

SISPA-prepared cDNA were submitted to the University of Wisconsin-Madison Biotechnology Center for sequencing on the Oxford Nanopore PromethION. Upon arrival, the PCR product concentrations were confirmed with the Qubit dsDNA high-sensitivity kit (Invitrogen). Libraries were prepared with up to 100-200 fmol of cDNA according to the Oxford Nanopore ligation-based sequencing kit SQKLSK109 and Native Barcoding kit EXP-NBD196. The quality of the finished libraries was assessed using an Agilent Tapestation (Agilent) and quantified again using the Qubit® dsDNA HS Assay Kit (Invitrogen). Samples were pooled and sequenced with an FLO-PRO002 (R9.4.1) flow cell on the Oxford Nanopore PromethION 24 for 72 hours. Data were basecalled using Oxford Nanopore’s Guppy software package (6.4.6) with the high accuracy basecalling model (read filtering parameters: minimum length 200 bp, minimum Qscore=9). Air sample AE0000100A8B3C was also sequenced on the Oxford Nanopore GridION to obtain a greater depth of coverage across all seven influenza C virus gene segments. A sequencing library was prepared for AE0000100A8B3C according to the Oxford Nanopore ligation-based sequencing kit SQK-LSK110 instructions. The sample was sequenced with an FLOMIN106 (R9.4) flow cell on the Oxford Nanopore GridION for 72 hours. Data were basecalled using Oxford Nanopore’s Guppy software package (6.4.6) with the high accuracy basecalling model (read filtering parameters: minimum length 20 bp, minimum Qscore=9).

### Sequencing analysis

Sequencing data generated from air samples were deposited in the Sequence Read Archive (SRA) under bioproject PRJNA950127. The removal of host reads was requested at the time of SRA submission using the Human Read Removal Tool (HRRT). The sequencing data were analyzed using a custom workflow. To ensure reproducibility and portability, we implemented the workflow in NextFlow and containerized all software dependencies with Docker. All workflow code and replication instructions are publicly available at (https://github.com/dholab/air-metagenomics). Briefly, the workflow starts by automatically pulling the study fastq files from SRA, though it has the option of merging locally stored demultiplexed fastq files as well. Then, reads are filtered to a minimum length (200bp) and quality score (Qscore=9), and adapters and barcodes are trimmed from the ends of the reads, all with the reformat.sh script in bbmap (39.01-0). The filtered fastq files for each air sample are then mapped to contaminant FASTA files containing common contaminants with minimap2 (v2.22). Reads that do not map to the contaminant FASTA files are retained and mapped to their sequencing run’s negative control reads to further remove contaminants present from library preparation. The cleaned fastq files for each air sample are then mapped to a RefSeq file containing human viruses downloaded from NCBI Virus using minimap2 (v2.22). The human virus reference file contains 835 viral genome sequences and was processed using the bbmask.sh command in bbmap (39.01-0) with default parameters to prevent false-positive mapping to repetitive regions in viral genomes. SAM files for each sample are converted to BAM format, again with reformat.sh. The workflow then completes by generating a pivot table of pathogen “hits,” which lists the number of reads supporting each mapped pathogen for each sample. For this study, we then imported the BAM alignments into Geneious Prime (2023.0.4) to inspect the mapping results visually. Genome coverage plots were created for several respiratory and enteric viruses detected in air samples using ggplot2 (3.4.1) with a custom R script (4.2.3) in RStudio (2023.03.0+386).

### Phylogenetic analysis

To compare the influenza C virus detected in the preschool air sample AE0000100A8B3C we downloaded 45 influenza C virus genome sequences for each of the seven gene segments from Genbank (HE, PB2, PB1, P3, NP, M, and NS). Accession numbers for each segment can be found in supplementary data 1. Consensus sequences were generated from AE0000100A8B3C with a minimum coverage of 20X. Sections with low coverage were masked with N and trimmed to the reference sequence length. Next, each set of influenza C virus gene segment sequences was aligned using MUSCLE (5.1) implemented in Geneious Prime (2023.0.4) with the PPP algorithm. We then used the Geneious Tree Builder (2023.0.4) to construct a phylogeny for each gene segment using the Neighbor-joining method and Tamura-Nei model with 100 bootstrapped replicates.

### SARS-CoV-2 RT-PCR

Air samples collected between December 2021 and May 2022 were tested for SARS-CoV-2 viral RNA using three different SARS-CoV-2 RT-PCR assays depending on their collection location as previously described ^19^. Air samples collected after May 2022 were tested for SARS-CoV-2 viral RNA using an RT-PCR protocol as previously described ^42^. Briefly, viral RNA was isolated from the air sample substrate using 300 μL of eluate and the Viral Total Nucleic Acid kit for the Maxwell 48 instrument (Promega), following the manufacturer’s instructions. RNA was eluted in 50 μL of nuclease-free water. Reverse transcription qPCR was performed using primer and probes from an assay developed by the Centers for Disease Control and Prevention to detect SARS-CoV-2 (N1 and N2 targets). The 20 μL reaction mix contained 5 μL of 4x TaqMan Fast Virus 1-Step Master Mix, 1.5 μL of N1 or N2 primer/probe mix (IDT), 5 μL of sample RNA, and 8.5 μL of nuclease-free water. The RT-PCR amplification was run on a LightCycler 96 at the following conditions: 37℃ for 2 minutes, 50℃ for 15 minutes, 95℃ for 2 minutes, 50 cycles of 95°C for 3 seconds and 55℃ for 30 seconds, and final cool down at 37℃ for 30 seconds. The data were analyzed in the LightCycler 96 software 1.1 using absolute quantification analysis. Air samples were called positive when N1 and N2 targets both had cycle threshold (Ct) values <40, inconclusive when only one target had Ct <40, and negative if both targets had Ct >40.

### Data Availability

The air sample sequencing data generated in this study have been deposited in the Sequence Read Archive (SRA) under bioproject PRJNA950127. The accession numbers for influenza C virus samples used in the phylogenetic analysis are provided in Supplementary Data 1.

## Code Availability

Code to replicate air sample sequencing analysis is available at https://github.com/dholab/air-metage-nomics.

## Supporting information

ICV Accession Numbers

## Data Availability

https://github.com/dholab/air-metagenomics

## Acknowledgments

This work was made possible by financial support through the National Institutes of Health grant (AAL4371). M.D.R. is supported by the National Institute of Allergy and Infectious Diseases of the National Institutes of Health under Award Number T32AI55397. The author(s) thank the University of Wisconsin Biotechnology Center DNA Sequencing Facility (Research Resource Identifier – RRID:SCR_017759) for providing PromethION sequencing services. We would like to acknowledge Eli O’Connor’s work in developing the iOS and Android Askidd mobile app to help streamline air sample metadata collection. We would like to thank all of the participating congregate settings for their partnership during this study.

## Author contributions

N.R.M contributed to the formal analysis, investigation, methodology, writing—original draft preparation and writing as well as revision—review and editing. M.D.R contributed to the conceptualization, data curation, formal analysis, investigation, methodology, project administration, visualization, writing—original draft preparation, writing—review and editing. D.H.O and S.L.O. contributed to the conceptualization, project administration, writing—original draft preparation, and writing—review and editing. M.R.S., O.E.H., A.A., W.C.V., M.J.B., and J.R.R. contributed to data curation, logistics, organization, and writing—review and editing. L.J.B. and M.T.A. contributed to data curation, resources, project management, and writing—review and editing. S.F.B, S.W., M.L., and M.M. contributed to logistics, organization, and writing—review and editing.

