## Supplemental Figure 1 for "Metagenomic sequencing detects human respiratory and enteric viruses in air samples collected from congregate settings"

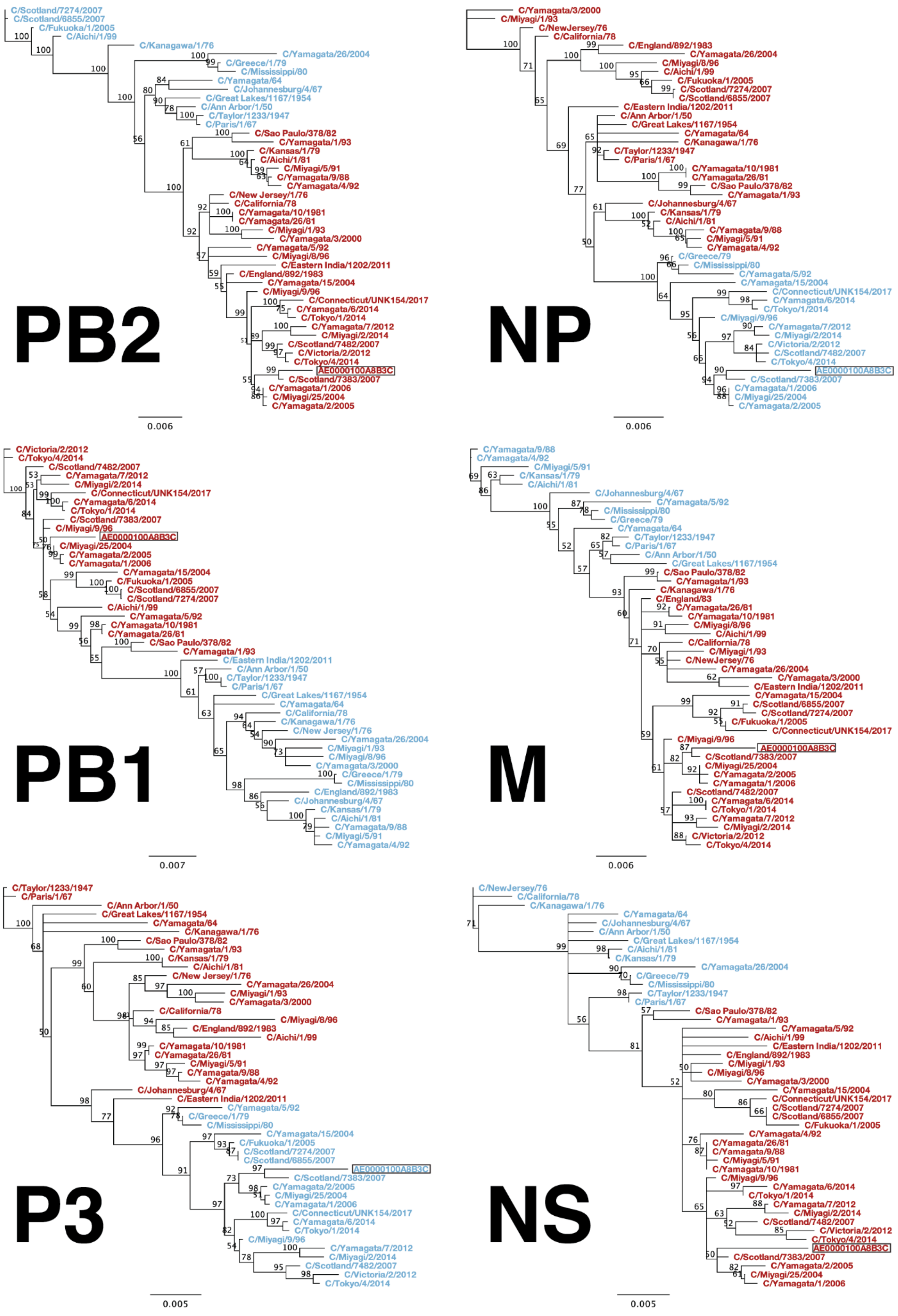
PB2
C/Scotland/7274/2007
C/Scotland/6855/2007
C/Fukuoka/1/2005
C/Aichi/1/99
C/Kanagawa/1/76
C/Yamagata/26/2004
C/Greece/1/79
C/Mississippi/80
C/Yamagata/64
C/Johannesburg/4/67
C/Great Lakes/1167/1954
C/Ann Arbor/1/50
C/Taylor/1233/1947
C/Paris/1/67
C/Sao Paulo/378/82
C/Yamagata/1/93
C/Kansas/1/79
C/Aichi/1/81
C/Miyagi/5/91
C/Yamagata/9/88
C/Yamagata/4/92
C/New Jersey/1/76
C/California/78
C/Yamagata/10/1981
C/Yamagata/26/81
C/Miyagi/1/93
C/Yamagata/3/2000
C/Yamagata/5/92
C/Miyagi/8/96
C/Eastern India/1202/2011
C/England/892/1983
C/Yamagata/15/2004
C/Miyagi/9/96
C/Connecticut/UNK154/2017
C/Yamagata/6/2014
C/Tokyo/1/2014
C/Yamagata/7/2012
C/Miyagi/2/2014
C/Scotland/7482/2007
C/Victoria/2/2012
C/Tokyo/4/2014
AE0000100A8B3C
C/Scotland/7383/2007
C/Yamagata/1/2006
C/Miyagi/25/2004
C/Yamagata/2/2005
0.006
NP
C/Yamagata/3/2000
C/Miyagi/1/93
C/NewJersey/76
C/California/78
C/England/892/1983
C/Yamagata/26/2004
C/Miyagi/8/96
C/Aichi/1/99
C/Fukuoka/1/2005
C/Scotland/7274/2007
C/Scotland/6855/2007
C/Eastern India/1202/2011
C/Ann Arbor/1/50
C/Great Lakes/1167/1954
C/Yamagata/64
C/Kanagawa/1/76
C/Taylor/1233/1947
C/Paris/1/67
C/Yamagata/10/1981
C/Yamagata/26/81
C/Sao Paulo/378/82
C/Yamagata/1/93
C/Johannesburg/4/67
C/Kansas/1/79
C/Aichi/1/81
C/Yamagata/9/88
C/Miyagi/5/91
C/Yamagata/4/92
C/Greece/79
C/Mississippi/80
C/Yamagata/5/92
C/Yamagata/15/2004
C/Connecticut/UNK154/2017
C/Yamagata/6/2014
C/Tokyo/1/2014
C/Miyagi/9/96
C/Yamagata/7/2012
C/Miyagi/2/2014
C/Victoria/2/2012
C/Scotland/7482/2007
C/Tokyo/4/2014
AE0000100A8B3C
C/Scotland/7383/2007
C/Yamagata/1/2006
C/Miyagi/25/2004
C/Yamagata/2/2005
0.006
PB1
C/Victoria/2/2012
C/Tokyo/4/2014
C/Scotland/7482/2007
C/Yamagata/7/2012
C/Miyagi/2/2014
C/Connecticut/UNK154/2017
C/Yamagata/6/2014
C/Tokyo/1/2014
C/Scotland/7383/2007
C/Miyagi/9/96
AE0000100A8B3C
C/Miyagi/25/2004
C/Yamagata/2/2005
C/Yamagata/1/2006
C/Yamagata/15/2004
C/Fukuoka/1/2005
C/Scotland/6855/2007
C/Scotland/7274/2007
C/Aichi/1/99
C/Yamagata/5/92
C/Yamagata/10/1981
C/Yamagata/26/81
C/Sao Paulo/378/82
C/Yamagata/1/93
C/Eastern India/1202/2011
C/Ann Arbor/1/50
C/Taylor/1233/1947
C/Paris/1/67
C/Great Lakes/1167/1954
C/Yamagata/64
C/California/78
C/Kanagawa/1/76
C/New Jersey/1/76
C/Yamagata/26/2004
C/Miyagi/1/93
C/Miyagi/8/96
C/Yamagata/3/2000
C/Greece/1/79
C/Mississippi/80
C/England/892/1983
C/Johannesburg/4/67
C/Kansas/1/79
C/Aichi/1/81
C/Yamagata/9/88
C/Miyagi/5/91
C/Yamagata/4/92
0.007
M
C/Yamagata/9/88
C/Yamagata/4/92
C/Miyagi/5/91
C/Kansas/1/79
C/Aichi/1/81
C/Johannesburg/4/67
C/Yamagata/5/92
C/Mississippi/80
C/Greece/79
C/Yamagata/64
C/Taylor/1233/1947
C/Paris/1/67
C/Ann Arbor/1/50
C/Great Lakes/1167/1954
C/Sao Paulo/378/82
C/Yamagata/1/93
C/Kanagawa/1/76
C/England/83
C/Yamagata/26/81
C/Yamagata/10/1981
C/Miyagi/8/96
C/Aichi/1/99
C/California/78
C/Miyagi/1/93
C/NewJersey/76
C/Yamagata/26/2004
C/Yamagata/3/2000
C/Eastern India/1202/2011
C/Yamagata/15/2004
C/Scotland/6855/2007
C/Scotland/7274/2007
C/Fukuoka/1/2005
C/Connecticut/UNK154/2017
C/Miyagi/9/96
AE0000100A8B3C
C/Scotland/7383/2007
C/Miyagi/25/2004
C/Yamagata/2/2005
C/Yamagata/1/2006
C/Scotland/7482/2007
C/Yamagata/6/2014
C/Tokyo/1/2014
C/Yamagata/7/2012
C/Miyagi/2/2014
C/Victoria/2/2012
C/Tokyo/4/2014
0.006
P3
C/Taylor/1233/1947
C/Paris/1/67
C/Ann Arbor/1/50
C/Great Lakes/1167/1954
C/Yamagata/64
C/Kanagawa/1/76
C/Sao Paulo/378/82
C/Yamagata/1/93
C/Kansas/1/79
C/Aichi/1/81
C/New Jersey/1/76
C/Yamagata/26/2004
C/Miyagi/1/93
C/Yamagata/3/2000
C/California/78
C/Miyagi/8/96
C/England/892/1983
C/Aichi/1/99
C/Yamagata/10/1981
C/Yamagata/26/81
C/Miyagi/5/91
C/Yamagata/9/88
C/Yamagata/4/92
C/Johannesburg/4/67
C/Eastern India/1202/2011
C/Yamagata/5/92
C/Greece/1/79
C/Mississippi/80
C/Yamagata/15/2004
C/Fukuoka/1/2005
C/Scotland/7274/2007
C/Scotland/6855/2007
AE0000100A8B3C
C/Scotland/7383/2007
C/Yamagata/2/2005
C/Miyagi/25/2004
C/Yamagata/1/2006
C/Connecticut/UNK154/2017
C/Yamagata/6/2014
C/Tokyo/1/2014
C/Miyagi/9/96
C/Yamagata/7/2012
C/Miyagi/2/2014
C/Scotland/7482/2007
C/Victoria/2/2012
C/Tokyo/4/2014
0.005
NS
C/NewJersey/76
C/California/78
C/Kanagawa/1/76
C/Yamagata/64
C/Johannesburg/4/67
C/Ann Arbor/1/50
C/Great Lakes/1167/1954
C/Aichi/1/81
C/Kansas/1/79
C/Yamagata/26/2004
C/Greece/79
C/Mississippi/80
C/Taylor/1233/1947
C/Paris/1/67
C/Sao Paulo/378/82
C/Yamagata/1/93
C/Yamagata/5/92
C/Aichi/1/99
C/Eastern India/1202/2011
C/England/892/1983
C/Miyagi/1/93
C/Miyagi/8/96
C/Yamagata/3/2000
C/Yamagata/15/2004
C/Connecticut/UNK154/2017
C/Scotland/7274/2007
C/Scotland/6855/2007
C/Fukuoka/1/2005
C/Yamagata/4/92
C/Yamagata/26/81
C/Yamagata/9/88
C/Miyagi/5/91
C/Yamagata/10/1981
C/Miyagi/9/96
C/Yamagata/6/2014
C/Tokyo/1/2014
C/Yamagata/7/2012
C/Miyagi/2/2014
C/Scotland/7482/2007
C/Victoria/2/2012
C/Tokyo/4/2014
AE0000100A8B3C
C/Scotland/7383/2007
C/Yamagata/2/2005
C/Miyagi/25/2004
C/Yamagata/1/2006
0.005

**Supplementary Figure 1. Phylogenetic analysis of influenza C virus six gene segments.** Phylogenetic trees for influenza C virus gene segments, including each of the genes encoding proteins for the polymerase complex (PB2, PB1, and P3), nucleoprotein (NP), matrix (M), and nonstructural protein (NS). Nucleotide sequences were aligned using MUSCLE (5.1). Phylogenies were constructed with the Geneious Tree Builder (2023.0.4) using the Neighbor-joining method and Tamura-Nei model with 100 bootstrapped replicates. Numbers above the branches indicate the bootstrap values with 100 replicates. ICV strain names are listed at the end of branches. ICV strains belonging to the C/Mississippi lineage are represented in light blue and C/Yamagata lineage in red.
